# Medicare Radiology Group Network Market Share: Recent Trends and Characteristics

**DOI:** 10.1101/2023.03.12.23287068

**Authors:** Ayis Pyrros, Brian Fornelli, Jorge Mario Rodríguez-Fernández, Stephen M. Borstelmann, Nasir Siddiqui, William Galanter

## Abstract

**Purpose:** Recent trends in US healthcare have seen growing consolidation of healthcare providers, including radiology groups, with fewer and larger radiology groups. We assessed recent trends and characteristics in radiology group network market share (NMS) across the US among Medicare beneficiaries.

**Methods:** Using freely available datasets CareSet DocGraph Hop Teaming, Medicare Physician Compare, and Medicare Physician and Other Supplier Public Use File (PUF), all radiologists were identified and associated to group practices annually between 2014 and 2017. Radiology groups outside the US, not present in all three databases, or with only one radiologist were excluded. The annual frequency of radiological exams performed was determined from the PUF and used to calculate the percentage of magnetic resonance and computed tomography (MR/CT) imaging performed as well as the number of beneficiaries undergoing MR/CT per radiology group. Physician referrers without evaluation and management codes for office visits in the PUF file were excluded from the DocGraph file. Provider connections were geospatially mapped and plotted. The percentage of radiology group NMS was calculated as the number of patients a group received, divided by the total number of potential and actual connections, and multiplied by 100. Univariate analysis of radiology group NMS was performed against a variety of characteristics and compared using Kruskal–Wallis and Dunn tests, as appropriate. Univariate linear regression was used to assess the association between NMS and calendar year, as well as average wait time and calendar year. Multivariate linear regression was used to model the radiology group cumulative normalized percentile NMS, with multiple predictors for the year 2017.

**Results:** Between 2014 and 2017, 1,764 unique radiology groups were identified, representing 17,879 radiologists in 2014, 18,143 radiologists in 2015, 20,915 radiologists in 2016, and 22,187 radiologists in 2017, with an average NMS per group for 2014 of 14.8%, 2015 of 14.5%, 2016 of 13.6%, and 2017 of 13.1%, demonstrating a small but statistically significant negative trend over four years (2014–2017), with a 0.6% decrease per year (P < 0.001). Average day weight across years (2014–2017) demonstrated a slight upward trend, with a 0.3% increase per year. The yearly percentage of MR/CT studies across all groups was 24.6–26.6%, with the most performed studies being chest radiography, mammography, and CT of the head without contrast. Univariate analysis of radiology group NMS was not significantly different between academic and nonacademic groups for all years (P > 0.05) but was significantly different for radiology-only versus multispecialty groups across all years (P < 0.05). Multivariate linear regression on 2017 data demonstrated statistically significant independent negative predictors for NMS including larger group size (50–99, >=100), higher practice MR/CT imaging percentage, and South location, while the increasing log of the number of beneficiaries undergoing MR/CT was a positive predictor, with an adjusted R-squared of 0.56 (all P < 0.01).

**Conclusions:** Among Medicare beneficiaries from 2014 to 2017, radiology group NMS (mean 14%, median 9.0%, IQR 4–19.2%), slightly decreased over time by 0.6% per year, despite occurring during a period of widespread practice consolidation. The negative predictors for NMS at the group level included larger group size, South region, increased average wait time, and higher MR/CT imaging percentage, while the positive predictor was the increased number of beneficiaries undergoing MR/CT. Of these predictors, radiology groups are most likely to increase NMS by decreasing average wait times.

**Highlights:**

1. Radiology group network market share (NMS) of Medicare beneficiaries is remarkably stable (mean 14%, median 9.0%, IQR 4–19.2%), with a statistically significant but small negative trend over four years (2014–2017), with a 0.6% increase per year.
2. Multivariate linear regression demonstrated that a higher percentage of magnetic resonance and computed tomography (MR/CT) imaging, group size (50–99, >= 100), South region, and longer average wait time are all negative predictors of NMS, and the only significant positive predictor of NMS is the increasing log number of beneficiaries undergoing MR/CT (all P < 0.05).
3. Of all predictors, average wait time is arguably the most malleable for a radiology group and should be closely monitored and reduced, as this may increase NMS. Additionally, having a diverse practice of services may offer additional opportunities to retain patients and grow NMS. In spite of radiology group consolidation, with the continued shift to value-based care, radiology groups need to be aware that the radiology market remains highly fragmented.
4. Source code for this project is available (https://github.com/apyrros/docgraph/) and can be freely run in Google Colab using Google Drive to store the necessary files.

## Introduction

Recent trends in US healthcare have seen growing consolidation of providers [1, 2], including radiology groups, with fewer and larger radiology groups [3]. Healthcare fragmentation is often cited as a driver of cost in the US healthcare system, with patients receiving fragmented care undergoing a comparatively higher number of diagnostic tests [4]. Patients receiving healthcare from multiple groups and systems may have diminished care coordination and increased cost, with healthcare systems suffering lost revenue and financial penalties in risk-shared contracts [5, 6, 7]. Reasons for fragmentation include a lack of specialist care, inadequate patient retention policies, poor provider communication, limited scheduling, patient distance, appointment availability, and health insurance policies [8]. Loss of patient market share can have significant financial consequences, with potential loss of revenue in the millions of dollars for healthcare systems [5, 6, 8]. Clearly not all patient sharing is undesirable: referrals to tertiary care centers are often necessary and desired to treat certain medical conditions efficiently. Nonetheless, medical group consolidation to achieve market share and reduce patient leakage is a commonly considered strategy in accountable care organizations (ACOs) to improve patient coordination and reduce redundant services [5].

Radiologists have a unique position in the medical space, serving the greatest number of individual Medicare beneficiaries as a specialty, effectively giving radiology the broadest reach into patients among physicians [9]. Because of this, the greater a radiology practice’s market share, the larger the reach into the group’s covered lives. There are numerous ways to consider market share. Here, we consider network market share (NMS), defined as patients seen divided by total actual and potential patients in that network. Accurate data on market share is challenging to source from third parties, and only the largest organizations may truly have a good understanding of their actual market share [5]. For smaller organizations, practice groups and smaller ACOs, it may be challenging to produce adequate market share data. The result is that many organizations and groups are blind to network market share, as only limited data, if any, is made available. Referral data is frequently shared with ACOs, such as a risk-shared insurance database [6], but detailed databases are not typically widely or publicly available. To assess NMS, we used the CareSet DocGraph Hop Teaming dataset [10, 11], which is based on administrative claims data from the Centers for Medicare and Medicaid Services (CMS), where at least 10 unique Medicare beneficiaries are shared between two healthcare providers connected in a directed graph structure, one of the largest graph databases freely available. Unfortunately, this dataset does not include details of exam types, diagnosis codes, and whether the shared patients were explicitly referred (i.e., directed to a radiology group). This type of data has been more appropriately described as a teaming dataset, showing how providers are connected and work together with common patients, rather than a referral set. Nonetheless, despite these limitations, the large volume of data provides insights into the US healthcare system and allows a deeper understanding of provider and patient connections.

There is a paucity of research on radiology group market share on the national level, even with the presumed prevalence and implications. Understanding the factors associated with radiology group NMS could potentially guide radiology groups and healthcare providers in process improvement and mitigation. Therefore, we aimed to assess recent trends and characteristics of radiology group market share across the US.

## Methods

### Datasets

Because this retrospective study used publicly available datasets without private identifiable information, it did not require Institutional Review Board approval. From the publicly available CMS Physician Compare database [12], we retrieved files for the fourth quarter of the available years (2014 through 2017), as previously chosen in other studies to maximize physician inclusion for the calendar year.

Radiologists were then identified using primary specialty codes for diagnostic radiology, interventional radiology, and nuclear medicine [3]. Also as previously described [2, 3], the Physician Compare underlying data source is Medicare’s Provider Enrollment, Chain, and Ownership System (PECOS), based on submitted claims data. This means that many providers are listed multiple times and affiliated to multiple group practices based on taxpayer identification numbers [3]. As has been previously done [3], radiologists with multiple practice affiliations were associated with the largest practice identified on the basis of the group size field in the Physician Compare dataset. The following features were then extracted for each radiologist identified: National Provider Identifier, organization name, organization practice ID, number of organization members, primary specialty, and state. Since we focused on radiology groups we did not exclude radiology trainees, i.e. recent medical school graduates, or part-time radiologists. For this study, we excluded radiologists who (1) were solo practitioners; (2) practiced outside the 50 United States and District of Columbia; or (3) were not present in the Physician Compare, Careset DocGraph Hop Teaming, or Medicare Public Use File (PUF) [13] databases.

For each year, we utilized the respective Careset DocGraph Hop Teaming file, which is based on administrative claims data, where at least 10 unique Medicare beneficiaries are shared between two healthcare providers and connected in a directed graph structure. The file details the number of unique patients shared between two providers, with the NPI numbers of the providers, and average wait time. The average wait time is the time it took, in days, for a patient to encounter the second provider after having seen the first provider. We then filtered the DocGraph file for only radiologists in the “to_npi” column of the database, utilizing the aforementioned NPI codes from the Physician Compare data.

Because the teaming file does not represent explicit referrals (i.e., patients specifically referred to radiology versus self-referrals or emergency visits), we filtered providers using Healthcare Common Procedure Coding System (HCPCS) evaluation and management codes (EM codes), starting with the prefix of 992 from the PUF for the same year. The reason for this is because radiologists can appear to refer patients to other providers in this dataset, when in reality patients are most likely simply returning to the ordering provider for additional management and follow-up. To mitigate this problem, we utilized the same year PUF data to exclude providers who do not utilize evaluation and management administrative code, providers such as diagnostic radiologists who do not have face-to-face patient encounters, similar to other studies [5]. This filter was generated using the unique beneficiary count from the PUF file to create a valid providers table, where providers had at least one or more EM codes.

NMS can be defined as the number of patients a radiology group receives divided by the total number of potential and actual patients in that network. For example, if a doctor sends 50 patients to his practice’s radiology group and 50 to an outside radiology group, the practice has 50% NMS from that doctor. At the radiology group level, if providers in a multispecialty group referred 1,000 patients to their practice’s radiology group and 2,000 patients to other radiology groups, the radiology group NMS would be 33%.

Using the structured query language (SQL) group by function we created a table of radiology groups, containing only one NPI number associated with the largest group. The same SQL methodology was used to associate referrer providers to radiology groups and sum the patient DocGraph counts. Likewise, we totaled the patient counts for each of these referring providers to any radiology group in the DocGraph database. We then calculated the radiology group NMS as the ratio of all patients the radiology group saw from all its associated providers to all patients seen by all associated providers regardless of which radiology practice they ultimately were associated with. Using the yearly PUF file we calculated the work relative value units (wRVUs) for each radiology group. We then calculated the percentage of magnetic resonance/computed tomography (MR/CT) studies performed at the group level using HCPCS code descriptions to generate a group weight for each radiology group.

### Practice Characteristics

As described in other studies [9, 14, 15], we grouped practices by the number of employed or affiliated radiologists: 2–9, 10–49, 50–99, and 100 or more; solo practices were excluded. Also as described in a prior study [14], practices whose names contained variants of either “radiology” or “imaging” were considered radiology-only; others were considered multispecialty.

As done in other studies [3] practices included in the Harvey L. Neiman Health Policy Institute Academic Radiology Practices list [16] and those whose names included variants of “univ,” “faculty,” “college,” or “school” were considered academic. The remaining practices were considered nonacademic. Each practice was also labeled geographically using US Census region labels (Midwest, Northeast, South, and West) [17].

Additional practice descriptions, based on the practice ZIP Code Tabulation Areas (ZCTA) from the Physician Compare file, were imputed, including the social deprivation index score and high needs score [18]. US Census data from 2015 [19] were utilized for area population, percent urban, and population, also based on a radiology group ZCTA from the Physician Compare file. To further characterize radiology groups by the work they performed, we calculated the percentage of MR/CT studies a group performed based on yearly PUF data. Additionally, we included the number of beneficiaries undergoing MR/CT.

Practice characteristics were compared using Pearon’s Chi-Square for categorical variables and t-tests for continuous variables. The NMS data distribution was assessed with Shapiro–Wilk test, and a P < 0.05 was considered significant. The Kruskal–Wallis test and the Dunn multiple comparison test were utilized as appropriate for nonparametric data.

A linear regression model was used to identify predictors of NMS for 2017, to avoid multi-year longitudinal clustering of radiology group data. Although other studies have utilized the year as a control variable, more advanced methods such as mixed-linear modeling or Generalized Estimating Equations are likely necessary [20]. Radiology group NMS was a very strongly left skewed variable and was normalized for all the groups in 2017. Therefore, the coefficients of the independent variables are associated with changes in percentage of group NMS relative to other groups. The specific practice variables utilized included practice size, radiology-only versus multispecialty practice, academic versus nonacademic practice, US Census region of the practice, average wait time, social deprivation index, Census percent urban, population, and radiology group wRVUs. We used the log of the group wRVUs and log of the number of beneficiaries undergoing MR/CT, because of severe left skewness in those data distributions, to improve the goodness of fit.

The files were processed utilizing python 3.7 (Wilmington, DE) and SQLite version 3.38 on Google Colab, with statistical analysis done using R (version 4) and Excel (Microsoft, Redmond, WA). A geographic plot of the group contections was generated in R, utilizing longitudinal and latitude data based on practice ZIP code. All tests of significance were evaluated as two-sided tests with α = 0.05.

## Results

Between 2014 and 2017, 1,764 unique radiology groups were identified, representing 17,879 radiologists in 2014, 18,143 radiologists in 2015, 20,915 radiologists in 2016, and 22,187 radiologists in 2017, (Table 1). A chi-square test of independence was performed to examine the relation between years and radiology-only practices, number of academic practices, demonstrating no relationship (P>0.05). The greatest number of groups were in the South region. The number of radiologists significantly increased between 2014 and 2017 (p=0.008), which would be expected from decreasing numbers of solo practices [3], and residency education cohorts being added to the pool of practicing radiologists. Excluded solo practices in this data numbered 286 in 2014, 268 in 2015, 271 in 2016, and 243 in 2017. We also saw average wait time for groups slightly increase over 2014–2017 (Table 1, P < 0.05). The mean radiology group NMS for 2014 was 14.8%, 2015 was 14.5%, 2016 was 13.6%, and 2017 was 13.1% (Table 1, Figure 1), with univariate linear regression showing a statistically significant (P < 0.001) but small negative trend over four years (2014–2017), a 0.6% decrease yearly. Univariate linear regression of average day weight across 2014–2017 demonstrated a slight upward trend, with a 0.3% increase per year (P < 0.001) (Figure 2).

**Table 1.** Baseline characteristics of radiology groups per year

| Characteristic <sup>1</sup> | 2014<br>(N = 1,203) | 2015<br>(N = 1,177) | 2016<br>(N = 1,211) | 2017<br>(N = 1,189) | P <sup>2</sup> |
| --- | --- | --- | --- | --- | --- |
| Number of radiologists, total | 17,879 | 18,143 | 20,915 | 22,187 | 0.008 |
| Radiology-only practice, vs multispeciality | 816 (68) | 789 (67) | 812 (67) | 802 (67) | 0.97 |
| Academic practice, vs nonacademic | 90 (7.5) | 92 (7.8) | 98 (8.1) | 99 (8.3) | 0.88 |
| Region |  |  |  |  |  |
| Midwest | 317 (26) | 309 (26) | 307 (25) | 280 (24) |  |
| Northeast | 296 (25) | 277 (24) | 316 (26) | 314 (26) |  |
| South | 363 (30) | 377 (32) | 357 (29) | 363 (31) |  |
| West | 227 (19) | 214 (18) | 231 (19) | 232 (20) |  |
| Percentage of market share by radiology group <sup>3</sup> | 14.8%, 9.5%<br>(4.3, 20.3) | 14.5%, 9.3%<br>(4.1, 14.5) | 13.6%, 8.8%<br>(4.0, 18.6) | 13.1%, 8.8%<br>(3.8, 18.3) |  |
| Average wait time (days) by radiology group <sup>3</sup> | 57.3, 57.9<br>(53.5, 61) | 57.6, 58.3<br>(53.9, 62.3) | 59.1, 58.4<br>(54.9, 62.9) | 58.1, 59.3<br>(54.6, 63.0) |  |
| Wait time >60 days | 428 (36) | 458 (39) | 532 (44) | 547 (46) | <0.001 |

<sup>1</sup> Data are given as number (percentage) for each group, unless otherwise specified.
<sup>2</sup> T-test; Pearson's Chi-squared test
<sup>3</sup> Mean, Median (quartile 1, quartile 3)

**Figure 1.**
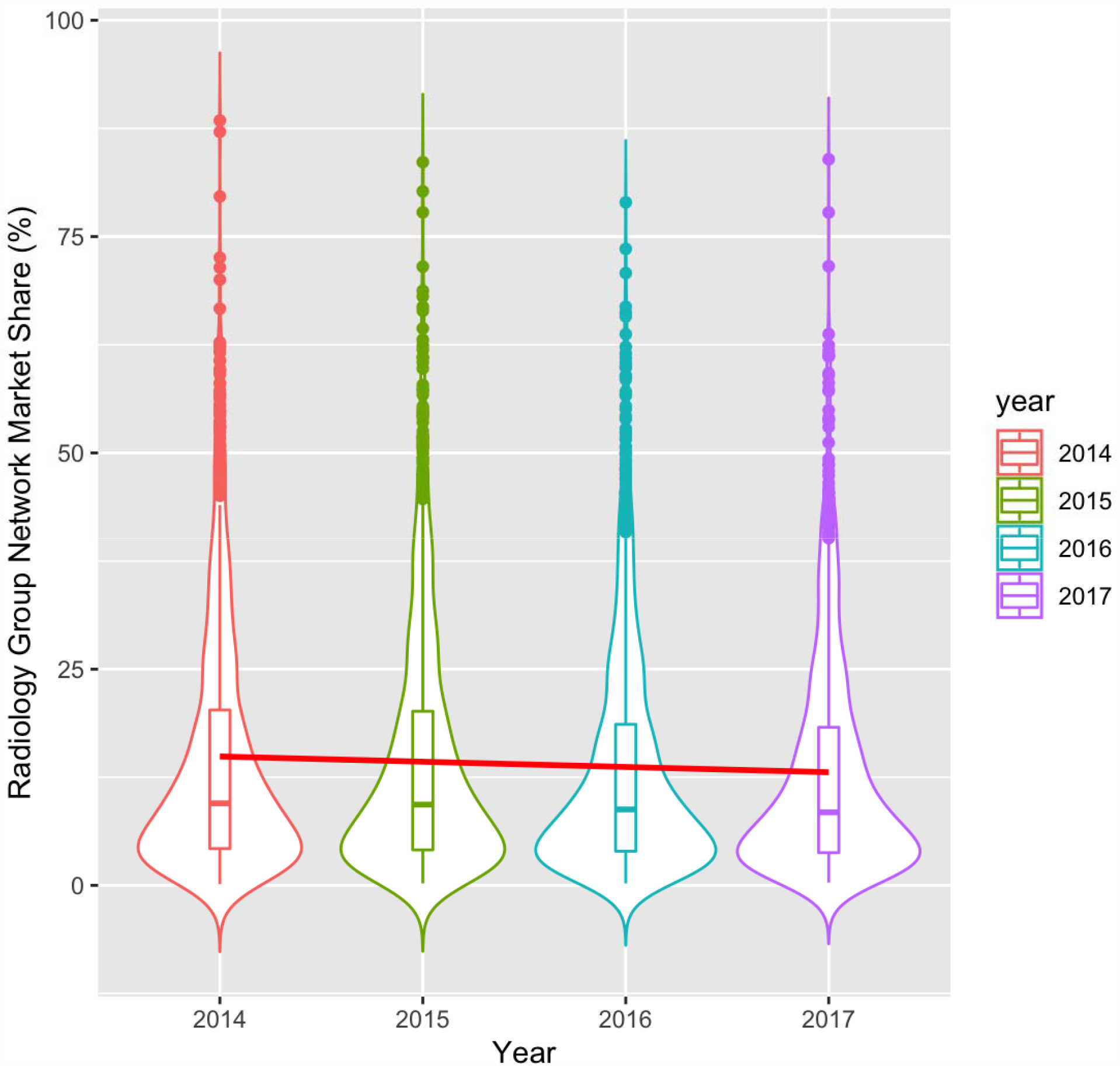
Radiology group network market share from 2014 to 2017. The violin plot of percent market share among all groups, with a trend line, shows a mild negative downward trend. Boxplots demonstrate the interquartile range and median (line).

**Figure 2.**
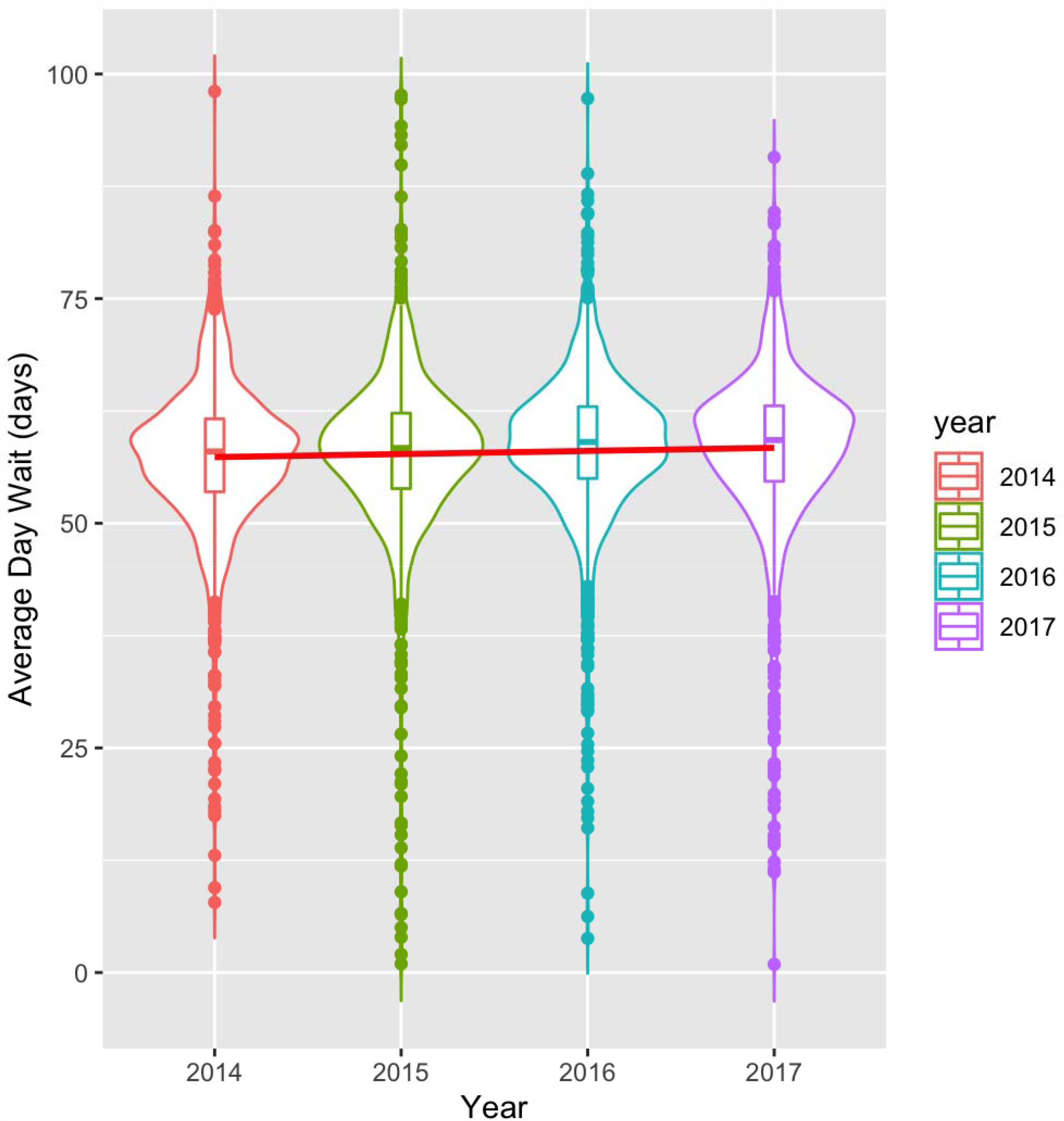
Average wait time among medicare radiology groups from 2014 to 2017. Boxplots demonstrate the interquartile range and median (line).

Table 2 shows the main distribution of radiology group NMS based on each group characteristic. Univariate analysis with the Kruskal–Wallis test shows that average group wait time >60 days vs <60 days was not statistically significant over 2015–2017, but was in 2014 (Table 2). Groups located in the West region tended to have a higher NMS compared to groups in the South and Northeast for 2014–2016 (Figure 3), with no significant difference across all groups in 2017, by Dunn’s test. Groups (50–99) had greater NMS (2–9, 10–49) across all years also by Dunn’s test, and larger groups (>=100) had greater NMS than smaller groups of 2–9 in 2016 and 2017. Academic versus nonacademic groups did not demonstrate a statistically significant difference in NMS (Table 2, Figure 4) between 2014 and 2017. Lastly, radiology-only groups had higher NMS (Table 2, Figure 5), which was significant across all years (P < 0.001).

**Table 2.**
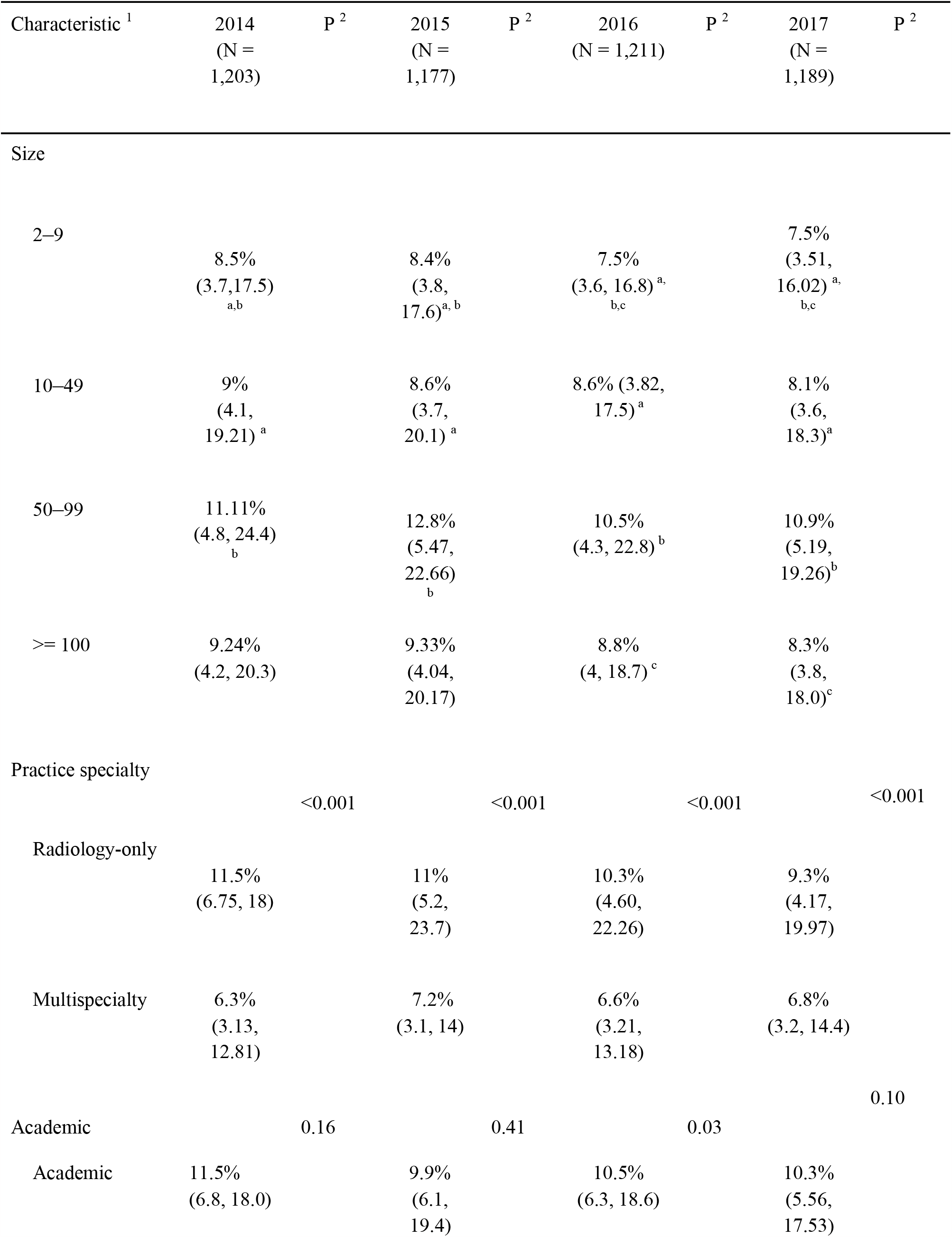
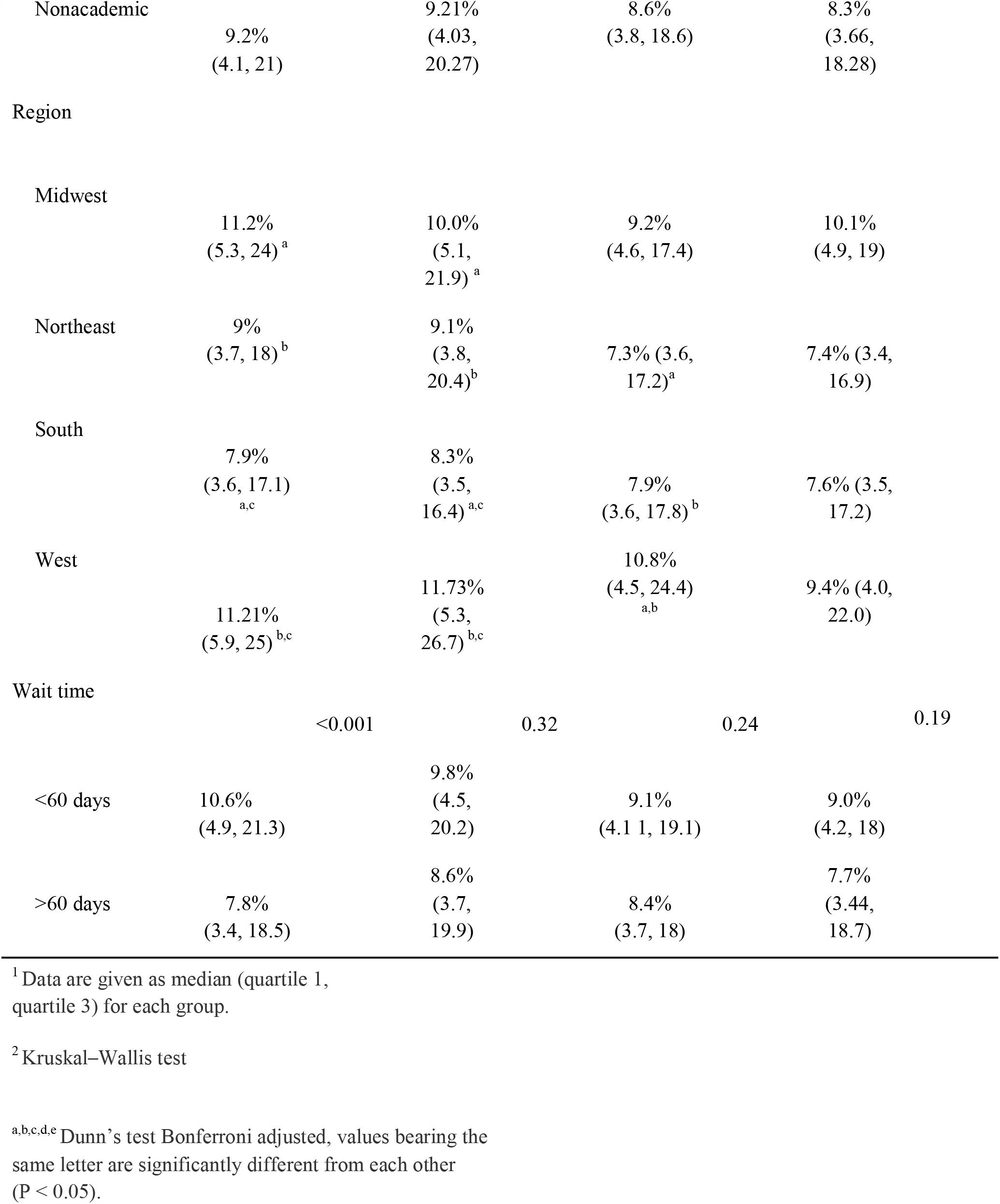
Yearly radiology group network market share by characteristic

**Figure 3.**
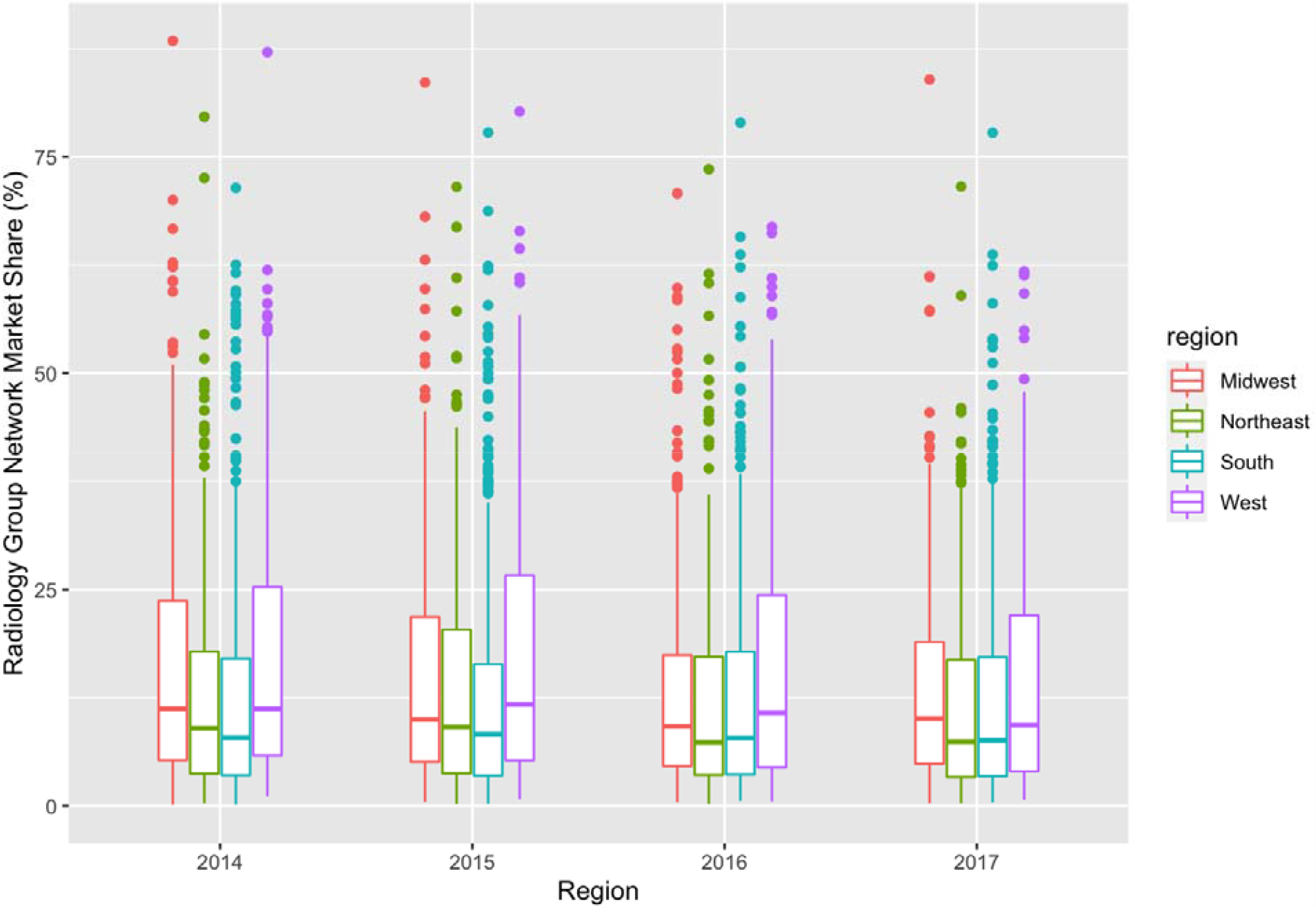
Radiology group network market share (percentage) from 2014 to 2017 across regions. Boxplots demonstrate the interquartile range and median (line).

**Figure 4.**
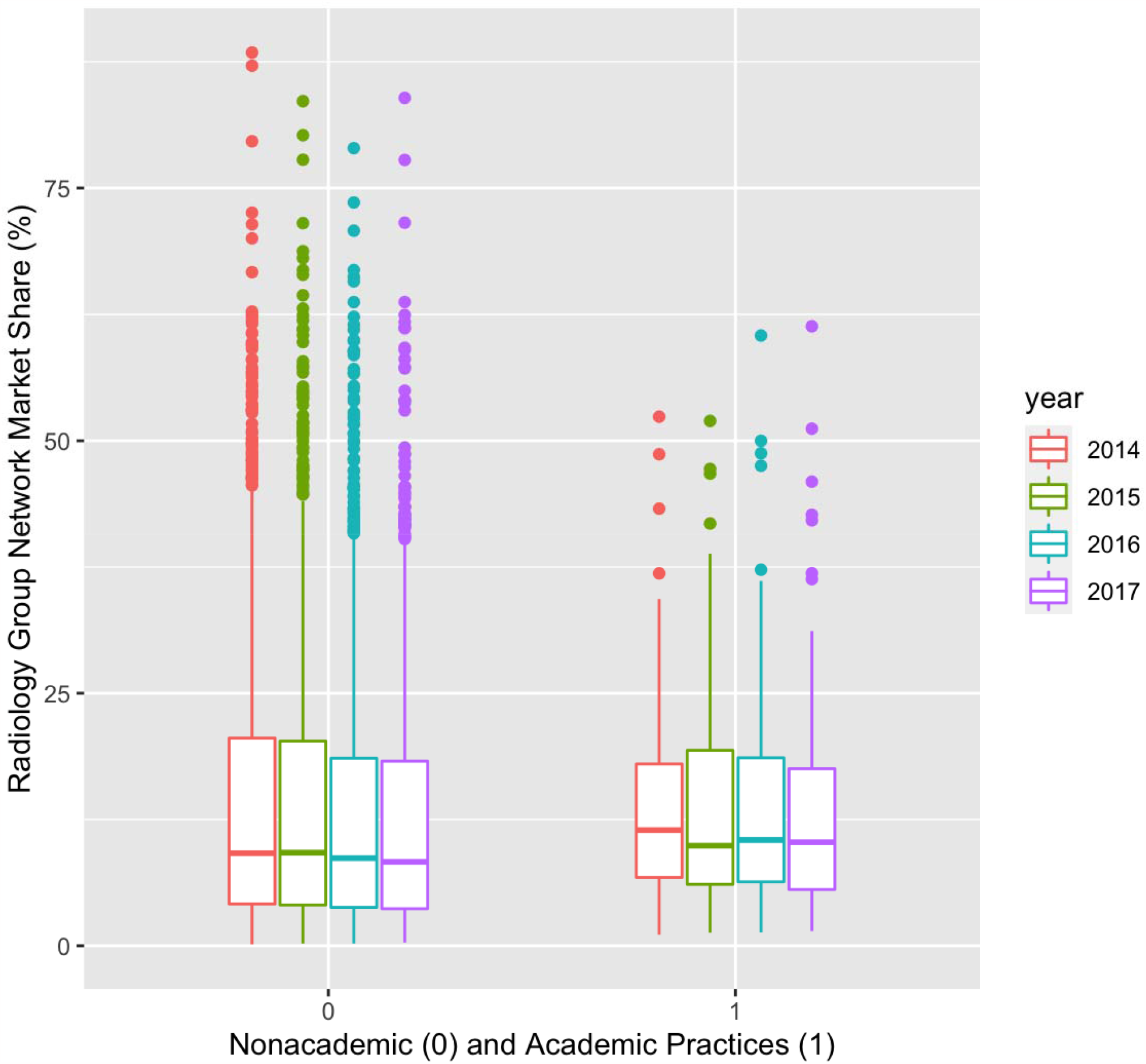
Radiology group market share (percentage) for nonacademic and academic practices across 2014 to 2017. Boxplots demonstrate the interquartile range and median (line).

**Figure 5.**
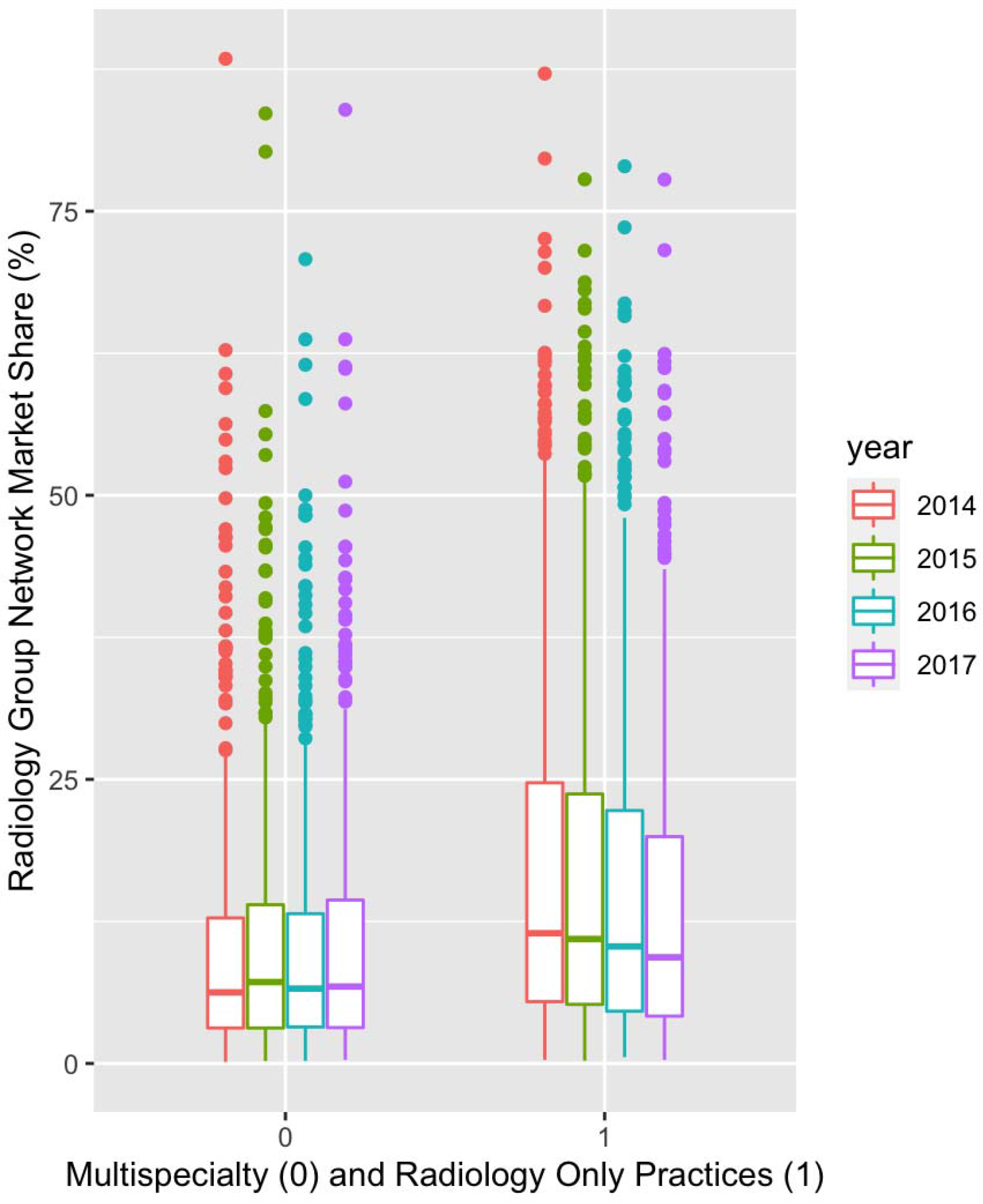
Radiology group network market share (percentage) for multispeciality and radiology-only groups across 2014 to 2017. Boxplots demonstrate the interquartile range and median (line).

Frequency of radiological exams are detailed in Table 3. Overall, chest radiographs (27.7–29%), mammograms (7.7–14.3%) and CT of the head without contrast (6.8–7.3%) were the most prevalent studies across 2014–2017. The yearly percentage of MR/CT studies across all groups was between 24.6– 26.6%.

**Table 3.**
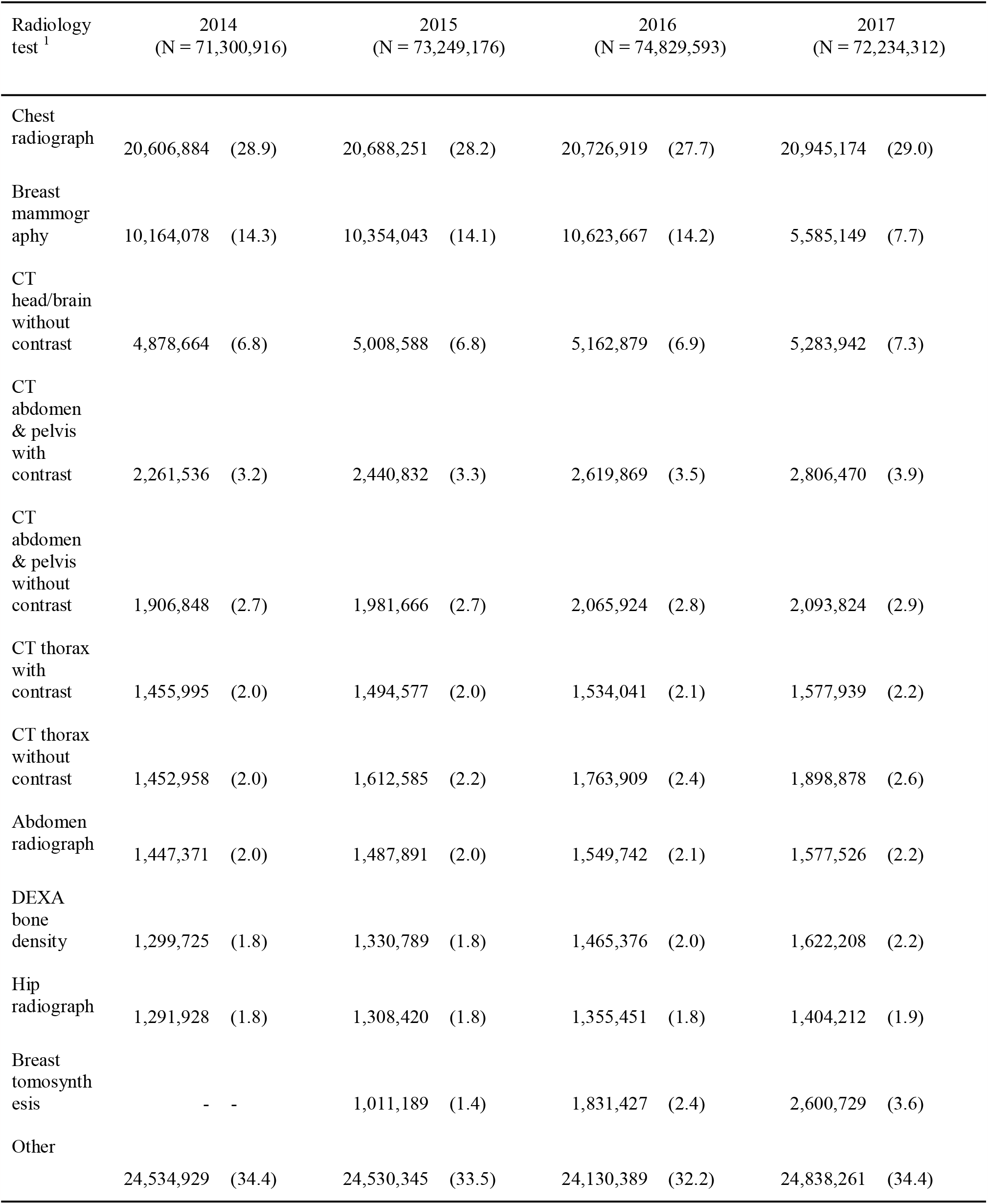
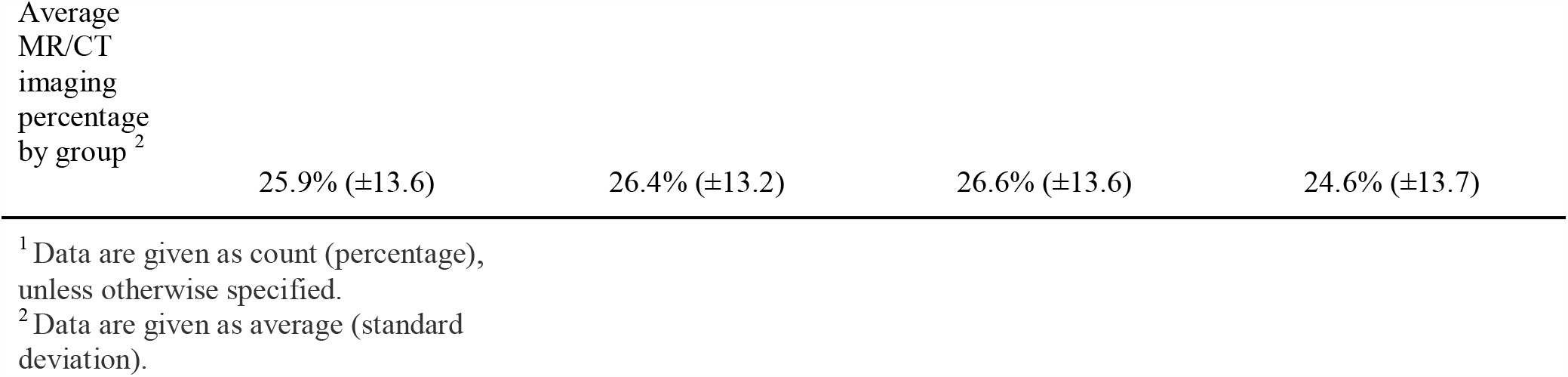
Imaging modality percentage seen by radiology groups per year in PUF

A graph of the 2017 radiology group referral network, representing 1,713,268 connections between providers and groups is shown in Figure 6. There are visibly less dense connections in the West region of the US. Conversely, there is marked density of healthcare providers and organizations in the East and South regions, which corresponds with US population density [21]. The majority of provider and radiology group connections have short distances, which are not seen because of size constraints [5].

**Figure 6.**
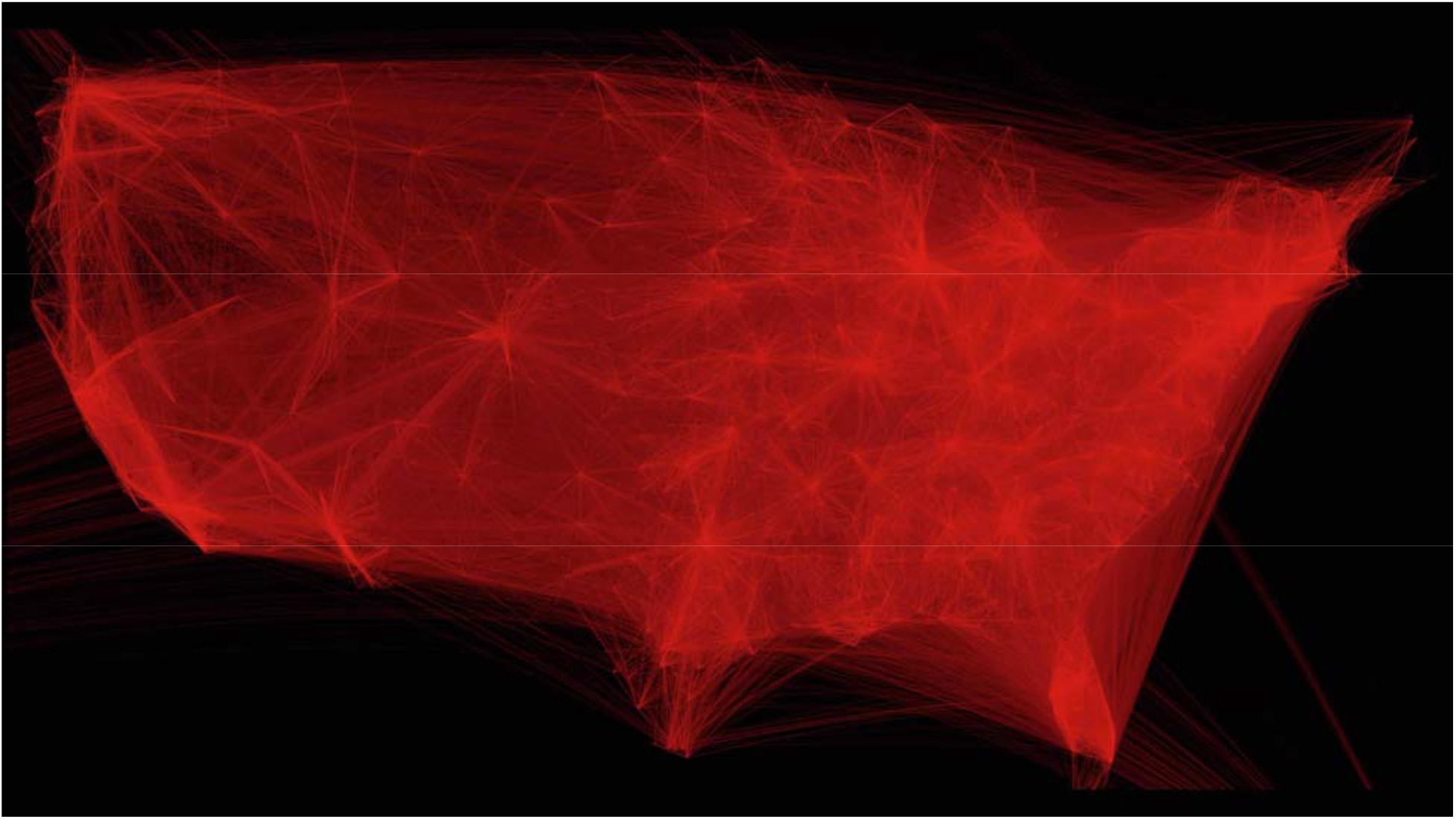
2017 radiology group referral network, representing 1,713,268 connections between providers and groups. The radiology network is geographically extensive, and mostly corresponding to population density.

Multivariate linear regression on the 2017 data (Table 4) demonstrated statistically significant independent negative predictors for network market share including group size (50–99, Beta = –0.124; 100 or more, –0.214), average wait time (Beta = –0.005), MR/CT imaging percentage (Beta = –0.724), and South location (Beta = 0.049), with the only positive predictor of network market share being the number of beneficiaries undergoing MR/CT (Beta = 0.131), with an adjusted R-squared of 0.56.

**Table 4.** Linear regression modeling of percentage network market share by group characteristics

| Characteristic | Coefficient | P value |
| --- | --- | --- |
| Size <sup>1</sup> |  |  |
|  | −0.003 | 0.850 |
| 10–49 |  |  |
|  | −0.124 | **<0.001** |
| 50–99 |  |  |
|  | −0.214 | **<0.001** |
| ≥100 |  |  |
|  | −0.008 | 0.119 |
| log(normalized wRVUs) |  |  |
|  | −0.035 | 0.132 |
| Academic center |  |  |
|  | −0.005 | **<0.001** |
| Average wait time |  |  |
|  | −0.030 | 0.273 |
| Percentage urban |  |  |
|  | −6.767e−07 | 0.070 |
| Population |  |  |
| Social deprivation index score | 2.696e−04 | 0.200 |
|  | 1.105e−05 | 0.957 |
| High needs score |  |  |
| Radiology group MR/CT percentage | −0.724 | **<0.001** |
| log(number of beneficiaries undergoing MR/CT) | 0.131 | **<0.001** |
| Region <sup>2</sup> |  |  |
|  | –0.008 | 0.601 |
| Northeast |  |  |
|  | –0.049 | **<0.002** |
| South |  |  |
|  | 0.025 | 0.148 |
| West |  |  |
| Radiology-only practice | 0.015 | 0.304 |
| F value | 95.93 |  |
| Adjusted R-squared | 0.562 |  |
| P value | <0.001 |  |

<sup>1</sup> Group size 2–9 was used as a reference.
<sup>2</sup> Midwest was used as a reference.

## Discussion

We found that the radiology group NMS was relatively constant with a slight downward trend over 2014– 2017, despite recent trends in radiology group consolidation, in this national study, which includes a large amount of mammography and radiography data. Although this data is pre-pandemic, we believe it still provides valuable insight into radiology networks and future trends.

Negative predictors for NMS were larger group size (50–99, >=100), increased average wait times, South region (versus Midwest), and increasing MR/CT imaging percentage. Groups may be best able to affect average wait times by ensuring adequate staffing and scheduling. Previous studies showed that using a SMART (*s*pecific, *m*easurable, *a*chievable, *r*elevant, and *t*ime-based) goal can effectively decrease wait times by more than 50% [22]. The average wait times we report based on the DocGraph data are derived from administrative data, and it is unclear how closely they reflect a patient’s actual date of service (i.e., date of exam), but we found there is a mild increasing trend in wait times. Similarly, other studies have demonstrated a trend of increasing wait times for new patients across different medical specialties, with large US metro area markets having an average wait of 32 days [23, 24]. Patient access to care remains a challenge across the spectrum of US healthcare [25], with causes attributed to increasing population, physician shortages, and administrative burdens [24].

As there is a continued shift towards value-based care and increased emphasis on care coordination, NMS will have negative financial consequences for organizations such as ACOs [26, 27]. However, seemingly little has changed in overall radiology group NMS among Medicare beneficiaries from 2014 to 2017, in part because patients remain largely unaware of the radiologist’s role or function [28]. Patients have little interaction with radiologists [28, 29] and therefore might not demonstrate a preference towards one radiology group over another. Instead, availability and location for radiological exams are the driving factors, primarily in the Medicare population, because individuals over 65 years, particularly those with complicated medical conditions, may not want to travel long distances [10, 30]. The physical location of a radiology group is not easily changed, but clearly has implications for NMS, and should be considered for new or expanding facilities.

The percentage of MR/CT imaging performed in a practice was a strong negative predictor of NMS, contrary to the number of beneficiaries undergoing MR/CT, which was a small positive predictor. This result means that practices with fewer absolute numbers of patients undergoing MR/CT would have smaller NMS. Counterintuitively, practices with a high percentage of MR/CT imaging also have smaller NMS, possibly indicating that they lack a diversity of services, such as mammography, sonography, DEXA bone density, or conventional radiography, or represent a teleradiology practice (Table 3). The recent trend toward larger radiology group sizes [3] does not appear to have impacted radiology group NMS. NMS relative to group size demonstrates that larger groups do not have a significant advantage yet, with an increasing number of provider connections. Also, larger groups may offer teleradiology services at multiple locations, which could decrease NMS at the group level [31]. Analysis by Census region demonstrates that groups in the South have less NMS, and the South region was a negative predictor of NMS. Additionally, there are more groups in the South relative to the West. In the West, there is a substantial dark area (Figure 6), which could represent differences in population density and the effect of a large self-contained integrated system in California, Oregon, and Washington [32].

Our study has several limitations, largely related to the available data formats. First, the data is entirely based on CMS claims, and therefore we cannot directly assess NMS in the private insurance market. Medicare patients, except for those in Medicare Advantage plans, typically have minimal or no restrictions on the doctor or facility from which they seek care. Therefore, we would expect more patient leakage and less NMS from traditional Medicare patients than insured lives covered by an ACO, or other entity engaged in value-based care. As a result, estimating market share from public Medicare data may well underestimate a group’s absolute market share. The fraction of explicit referrals and the referral reasons are unknown; in other words, we do not know whether patients chose a facility and group on their own or were directed by their physician to a specific radiology practice. We also realize that several groups may be contained within a single system, and in such cases, NMS could be calculated differently. Likewise, some radiologists might belong to more than one group (part-time, locums) and some groups may have a presence in multiple states, which could affect NMS, such as a teleradiology group. In addition, it cannot be determined how much of the wait is due to the group’s scheduled availability and how much was the patient waiting to schedule. A particular example is screening mammography, which does not require a physician order [33]; therefore, provider-to-provider wait times may have no relationship to the timing of the exam performed.

In conclusion, among CMS beneficiaries, while radiology group NMS demonstrates a slightly negative trend from 2014 to 2017, the degree of this decline is surprisingly underwhelming when compared to the extent of market consolidation during the same period. For the greater part, there was no measurable increase in the radiology group NMS. The US healthcare system remains highly fragmented, despite healthcare system consolidation, which can impede care [34]. For multimodality radiology practices, the most actionable metric to improve network market share is suggested to be average wait time. Practice leaders and/or management may wish to focus on accurate monitoring and optimization of this factor to improve NMS.

The source code is provided freely at https://github.com/apyrros/docgraph/ and can be used on Google Colab.

## Data Availability

All data produced in the present study are available upon reasonable request to the authors

https://github.com/apyrros/docgraph/

## Acknowledgements

The authors would like to thank Monica Harrington for editing and reviewing this manuscript.

